# Novel 10-channel phased-array coil design for carotid wall MRI at 3T

**DOI:** 10.1101/2022.12.01.22283007

**Authors:** Matthijs H.S. de Buck, Peter Jezzard, Robert Frost, Chris Randell, Katherine Hurst, Robin P. Choudhury, Matthew D. Robson, Luca Biasiolli

**Author notes:** **Corresponding author:** Luca Biasiolli.

## Abstract

**Background:** Accurate assessment of plaque accumulation near the carotid bifurcation is important for the effective prevention and treatment of stroke. However, vessel and plaque delineation using MRI can be limited by low contrast-to-noise ratio (CNR) and long acquisition times. In this work, a novel 10-channel phased-array receive coil design for bilateral imaging of the carotid bifurcation using 3T MRI is proposed.

**Methods:** The novel 10-channel receive coil was compared to a commercial 4-channel receive coil configuration using data acquired from phantoms and healthy volunteers (N = 9). The relative performance of the coils was assessed, by comparing signal-to-noise ratio (SNR), g-factor noise amplification, and the CNR between vessel wall and lumen using black-blood sequences. Patient data were acquired from 12 atherosclerotic carotid artery disease patients.

**Results:** The 10-channel coil consistently provided substantially increased SNR in phantoms (+88 ± 2%) and improved CNR in healthy carotid arteries (+62 ± 11%), or reduced g-factor noise amplification. Patient data showed excellent delineation of atherosclerotic plaque along the length of the carotid bifurcation using the 10-channel coil.

**Conclusions:** The proposed 10-channel coil design allows for improved visualization of the carotid arteries and the carotid bifurcation and increased parallel imaging acceleration factors.

## 1. Introduction

Atherosclerosis in the carotid arteries is one of the leading causes of stroke^1–4^, with the majority of plaque accumulation occurring near the carotid bifurcation. Accurate assessment of the size, shape, location, and composition of atherosclerotic plaques^1,3,5^ is important for the effective diagnosis and treatment of the disease and the prevention of ischaemic events.

Magnetic resonance imaging (MRI) can be used for non-invasive *in vivo* characterization of atherosclerotic plaque in the carotid arteries^5–7^. The different contrast weightings in MRI facilitate a comprehensive characterization of the vessel wall and the plaque^8–11^, as well as visualization of the arterial blood flow. Accurate MRI assessment of plaque size and composition, which are indicative of plaque vulnerability^2,12^, is constrained by the carotid image resolution and signal-to-noise ratio (SNR) that can be achieved within a clinically reasonable scan time.

Moreover, plaque lipid can be accurately quantified by T2 mapping on a voxel-by-voxel basis, as demonstrated in endarterectomy patients by histological validation ^7,11,13,14^. This MRI technique can be used to study the relationship between plaque lipid content and symptomatic status, and to identify patients at higher risk of plaque rupture. However, it requires sufficient SNR in multiple spin echo images at different echo times to generate robust T2 estimates for each plaque voxel, thus it would clearly benefit from increased coil sensitivity at carotid depth.

The carotid bifurcation is located in a relatively superficial part of the neck, at a typical depth of 3 cm below the skin^15–17^, albeit deeper in overweight patients, who are at higher risk of atherosclerotic complications. Both the longitudinal location (here longitudinal is defined as the location along the vessel in the head-foot direction) and the depth of the carotid bifurcation can vary substantially among subjects due to physical differences in neck and vascular anatomy. This means that an effective MR receive coil for imaging near the carotid bifurcation requires high SNR at a sufficiently large penetration depth and longitudinal coverage in order to accommodate a wide range of anatomies.

In addition to high SNR, accurate carotid plaque characterization requires high-resolution images to accurately visualize the detailed (<0.5 mm) features of the plaque composition. Parallel imaging techniques^18^ are often used to acquire data at high resolutions with reduced scan times, at the cost of a loss in SNR. The relative loss of SNR can further degrade depending on the coil geometry being used^19^, so coil configurations which provide low amounts of noise amplification at high acceleration factors are desirable for carotid MRI^20,21^.

The advantages of phased-array coils for carotid artery imaging have been established by Hayes et al.^22^. For imaging near the carotid bifurcation at 3T, which is recommended over 1.5T in clinical practice because of its increased SNR^10^ and high clinical availability, various studies into the optimal coil configurations are available^15,17,20,23–26^. Those studies at 3T use between 4 and 16 coil channels for bilateral imaging. 30-channel coils for carotid MRI have been shown to facilitate high parallel imaging acceleration factors, but with limited SNR penetration^20,27^.

Coil configurations consisting of few but large receive elements typically benefit from large spatial coverage, but with limited SNR^17,20^. Increased numbers of small coil channels can provide improved superficial SNR, but with reduced penetration depth and flexibility^20,26^. In this work, a new 10-channel coil configuration for accurate bilateral visualization of the carotid bifurcation using parallel imaging acceleration is proposed. The achieved SNR, noise amplification, and vessel visualization using this coil is compared to results obtained using a commercial 4-channel carotid coil for phantom and *in-vivo* acquisitions. In a recent paper, Zhang et al.^20^ compare the performance of three different (6-, 8-, and 30-channel) carotid coil designs to the performance of the same commercial 4-channel coil which is used in this study. Therefore, the performance of the 10-channel coil proposed here relative to the 4-channel coil can be compared to their results to put the performance of the 10-channel coil into the context of those other designs.

## 2. Methods

### 2.1 Coil design

All data were acquired using a newly developed 10-channel phased-array receive coil (PulseTeq, Chobham, United Kingdom) and compared to results obtained from a widely used commercial 4-channel phased-array receive coil (MachNet BV, Roden, The Netherlands). The measurement setups using both coils are shown in Figure 1. Both coils were designed for bilateral imaging of the carotid arteries near the carotid bifurcation.

**Figure 1:**
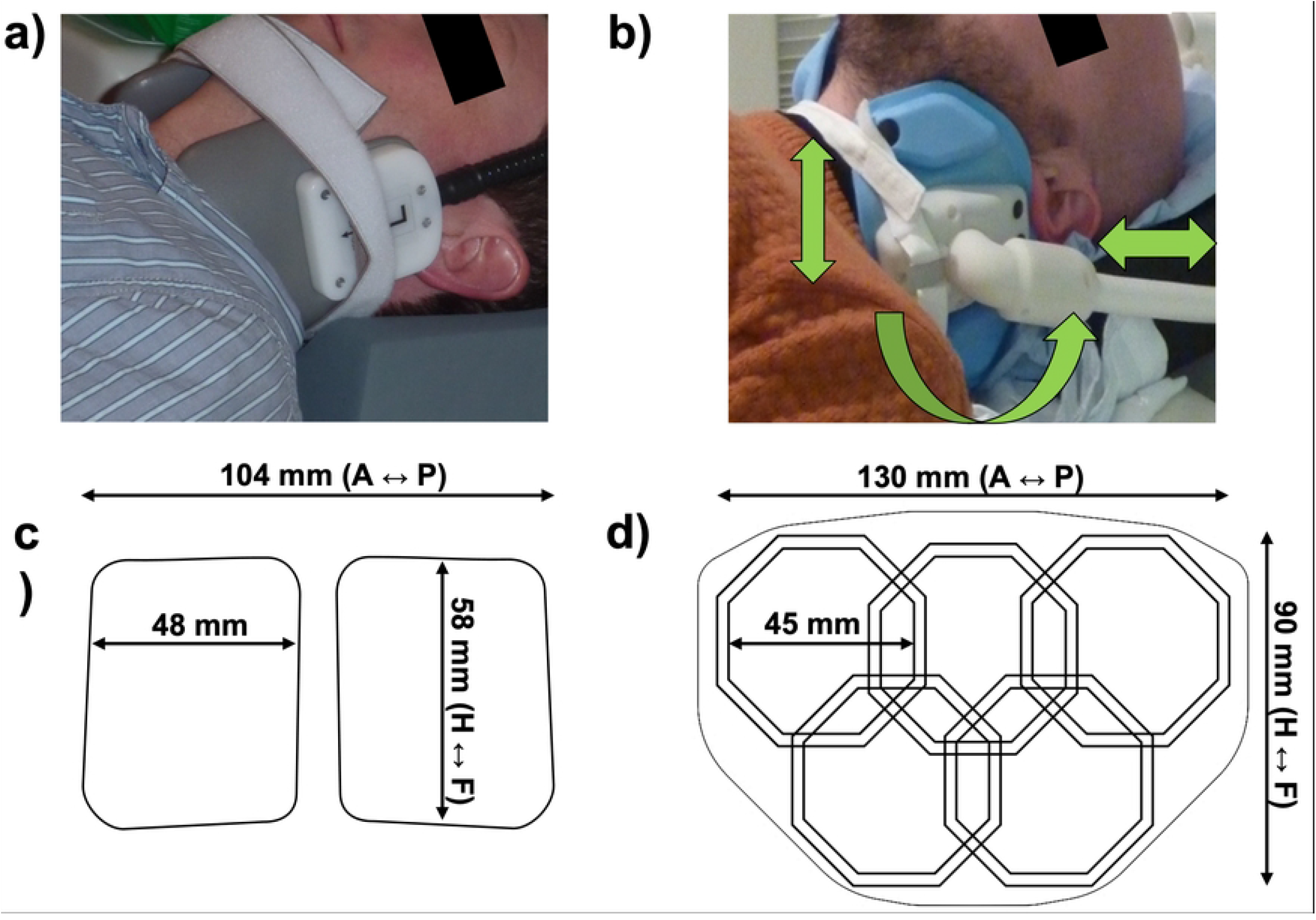
The two coils used in this work. **(a)**: The 4-channel coil positioned around the neck of a volunteer. **(b):** The 10-channel coil positioned around the neck of a (different) volunteer. The green arrows indicate the degrees of freedom of the coil positioning around the neck of the subject. **(c-d)** Relative positions and dimensions of 4-channel^20^ and 10-channel coils and their individual channels (figures show one of the bilateral sides).

The 4-channel coil consists of two bilateral sets of paired transverse channels. The 10-channel surface coil consists of two sets of octagonal elements with custom-made low-impedance preamplifiers, positioned in an ‘Olympic ring’-configuration (Figure 1c). Each side of the coil (overall dimension = 130 × 90 mm) is made of 5 elements of size = 45 mm with both active and passive protection. The coils were designed to be flexible and adaptable to different neck sizes, and are mounted on support arms that can bend and rotate for improved positioning with respect to the carotid bifurcation while maintaining high patient comfort. The coils are surrounded by foam covers to ensure optimal patient comfort and adequate isolation.

### 2.2 Phantom study

Data from a cylindrical short T1 phantom (15 cm diameter) were acquired for quantitative comparison of the relative performance of the two coils.

A single-slice spin-echo sequence was used for SNR measurements (TR/TE = 300ms/10ms, resolution 1.2×1.2×3.0mm, matrix size 256×256, total scan time 3:32 minutes). The same slice was scanned 6 times with 10 second pauses for temporal SNR (tSNR) calculations, assuming negligible motion and scanner drift.

Multi-slice T1-weighted turbo-spin-echo data were acquired for estimation of phantom g-factor maps of both coils in both coronal (10 slices) and transverse (20 slices) scan orientations. Sequence parameters: TR/TE = 1000/13ms, resolution 0.9×0.9×2.0mm, in-plane matrix size 256×256, 100% slice gap, turbo factor 10. Total scan time was 1:21 and 1:50 minutes for the coronal and transverse orientations, respectively.

### 2.3 *In vivo* study

Nine healthy volunteers (mean ± SD in age and weight: 33.2 ± 7.0 years; 78 ± 5 kg) were imaged with both the 4- and 10-channel coils on a Siemens (Erlangen, Germany) Verio 3T scanner using the DANTE-MESE sequence^11,28^ to acquire 5 transverse slices over 10 cm centered at the carotid bifurcation and estimate T2 maps of the arterial wall. The following Multi-Echo Spin-Echo (MESE) acquisition parameters were used: 14 echoes (TE = 9.1 to 127.4 ms), TR = 2000 ms, FOV = 128 × 128 × 100 mm, matrix size = 192 × 192, voxel size 0.67 × 0.67 × 2 mm, slice gap = 100%, scan time 4 min. A Delay Alternating with Nutation for Tailored Excitation (DANTE^29^) preparation before each readout was used for flowing blood signal suppression. The following DANTE parameters were used: gradient amplitude = 18 mT/m, 120 RF pulses, flip angle = 8°, RF pulse interval = 500 μs. Bright-blood Time-of-Flight (TOF) MR Angiography (MRA) was acquired to localize carotid bifurcations. Additional multi-slice T1-weighted turbo-spin-echo data were acquired in one healthy volunteer for the estimation of *in vivo* g-factor maps. 12 transverse slices were acquired with TR/TE = 1090ms/13.1ms, resolution 0.6 × 0.6 × 2.0 mm, in-plane matrix size 256×252, 100% slice gap, turbo factor 7, total scan time 2:41 minutes. Data were acquired under an agreed technical development protocol approved by the Oxford University Clinical Trials and Research Governance office, in accordance with International Electrotechnical Commission and UK Health Protection Agency guidelines.

12 patients with atherosclerosis (72.3 ± 9.4 years, 80.6 ± 11.7 kg) were scanned at carotid plaque locations using slices perpendicular to the direction of the vessel with the 10-channel coil using DANTE-FSE (Fast Spin Echo) T1-weighted imaging. FSE acquisition parameters were TR = 1280 ms, TE = 13 ms, FOV = 150 × 150 mm, matrix size 256×256 (0.59 × 0.59 mm resolution), echo train length = 7, slice thickness = 2 mm, slice gap = 100%, scan time ∼ 3 min). DANTE preparation parameters were gradient amplitude = 18 mT/m, 64 RF pulses, flip angle = 8°, RF pulse interval = 1 ms. A bright-blood TOF MRA was acquired to localize carotid bifurcations and lumen stenoses. 13 interleaved T1-weighted slices were acquired at the level of the atherosclerotic plaques (affected carotid side based on Doppler Ultrasound). Ethical approval was obtained from the UK National Research Ethics Services and patients provided written informed consent.

### 2.4 Image analysis

In the phantom study, SNR was calculated for single spin-echo acquisitions. For each pixel, the SNR was calculated as

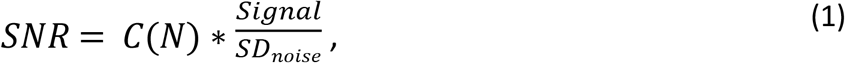

using the signal standard deviation in a 50 × 50 pixel background region-of-interest (without visible artifacts, e.g. ghosting/Gibbs ringing) as a noise reference (*SD*_*noise*_)^30^. *C*(*N*) is the SNR correction factor based on the number of coil channels as described by Gilbert^31^, which gives 0.695 for the 4-channel coil and 0.703 for the 10-channel coil. The temporal stability of the two coils was compared using the tSNR, calculated based on the pixel-wise signal mean and standard deviation of the 6 consecutively acquired slices.

In the *in vivo* study, wall/lumen contrast-to-noise ratio (CNR) for the healthy volunteers was estimated as the SNR difference between the carotid vessel wall and its lumen. Inner and outer vessel wall boundaries were segmented following published procedures^11^. Resulting CNR values were compared for images acquired using the 10-channel and the 4-channel coil. For all nine volunteers, results were compared at each of the 14 different echo times.

Since images were acquired with the 10-channel and 4-channel coil at different times (scan-rescan during the same session), identical voxel locations could not be assumed for quantitative statistical analysis. Therefore, we tested the null hypothesis that data were independent random samples drawn from the same normal distribution, using a two-sample t-test at 5% significance level.

Voxel-wise T2 values were estimated in the carotid wall by fitting an exponential decay curve to the signal intensity of the 14 echoes using a Levenberg-Marquardt nonlinear least squares algorithm^11^. T2 maps were generated using data acquired with the 10-channel and the 4-channel coil. Statistical comparisons were performed for the estimated T2 values of the healthy vessel wall and their standard errors using a two-sample t-test.

The geometry factor (g-factor) noise amplification metric is used to assess the parallel imaging performance of a receiver coil when using methods such as SENSE^19^ or GRAPPA^32^. For an acceleration factor of R, the reconstructed SNR (*SNR*_*PI*_) is given by

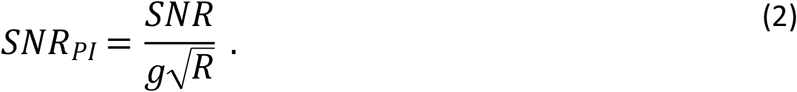

The spatially variant g-factor noise amplification of both coils was estimated for the (phantom and *in vivo*) turbo-spin-echo acquisitions at retrospectively undersampled acceleration factors of 2 and 3. For this, the method described by Breuer et al.^33^ was used to calculate the g-factors after application of different GRAPPA kernels, using a calibration region of 32 × 32 k-space points. Analysis was performed using Matlab R2019a (MathWorks, Natick, MA).

## 3. Results

### 3.1 Phantom comparisons

SNR profiles for both the 4-channel coil and the 10-channel coil in a single-slice spin-echo acquisition are shown in Figure 2. The 10-channel coil consistently gives an increased SNR relative to the 4-channel coil, with higher increases closer to the edge of the phantom in the left-right direction and closer to the centre of the coils in the longitudinal direction. The mean (± standard deviation of mean) SNR gain along the longitudinal direction shown in Figure 2b is 88 ± 2%, with the largest SNR gain (>100%) at around 5 cm above and below the center. The tSNR over the same region also increased significantly (p < 0.001) for the 10-channel coil. At a depth of 3 cm, the average tSNR increased by 80 ± 8% for the 10-channel coil (data not shown).

**Figure 2:**
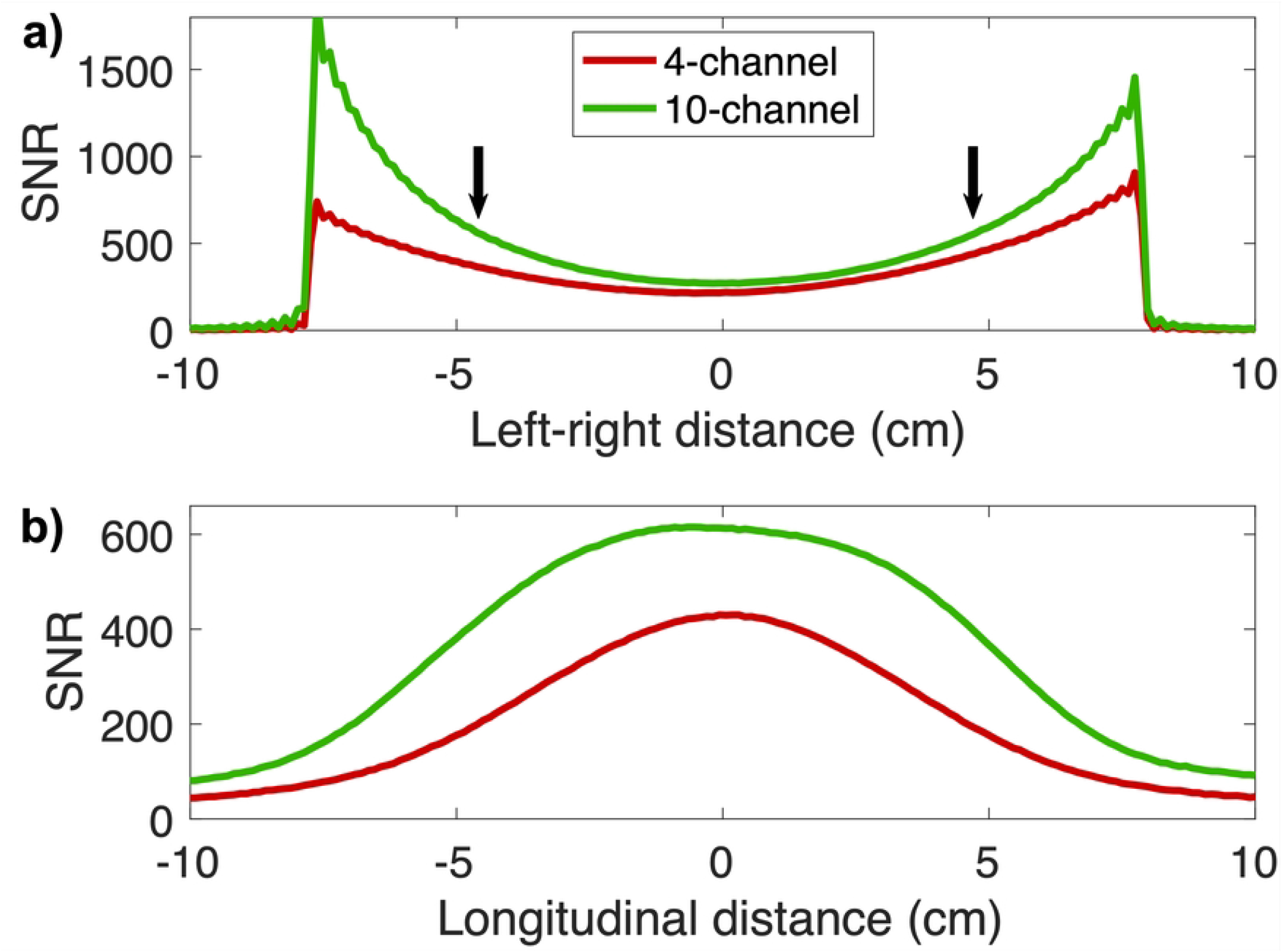
SNR profile of the two coils in a phantom, based on coronal single-slice acquisitions. **(a):** SNR in the left-right direction, along the line with the highest SNR for each coil. **(b):** Mean SNR in the longitudinal direction at 3 cm from the edge of the phantom at both sides (corresponding to the black arrows in (a)), which corresponds to the approximate typical depth of the carotid bifurcation.

Figure 3 shows the estimated g-factor noise amplification in the phantom using both coils at R = 2 (Figure 3a) and R = 3 (Figure 3b). The two top rows show examples of the reconstructed slices and g-factor distributions in the phantom in a single coronal slice for all four cases (both coils and both GRAPPA acceleration factors). The bottom row shows the maximum noise amplification values of all coronal and transverse slices. For coronal acquisitions, the 10-channel coil consistently achieves a significant (p < 0.001) g-factor reduction of 47 ± 7% at R = 2, and of 58 ± 3% at R = 3. For transverse acquisitions, the noise amplification is lower for both coils, probably due to the combination of smaller in-plane size of the phantom and increased spatial separation of receive channels in transverse acquisitions. Lower g-factors are visible for the 10-channel coil for some of the off-centre transverse slices where the 4-channel coil has an increased maximum g-factor. However, no overall statistically significant difference is observed between the two coils for the transverse acquisitions.

**Figure 3:**
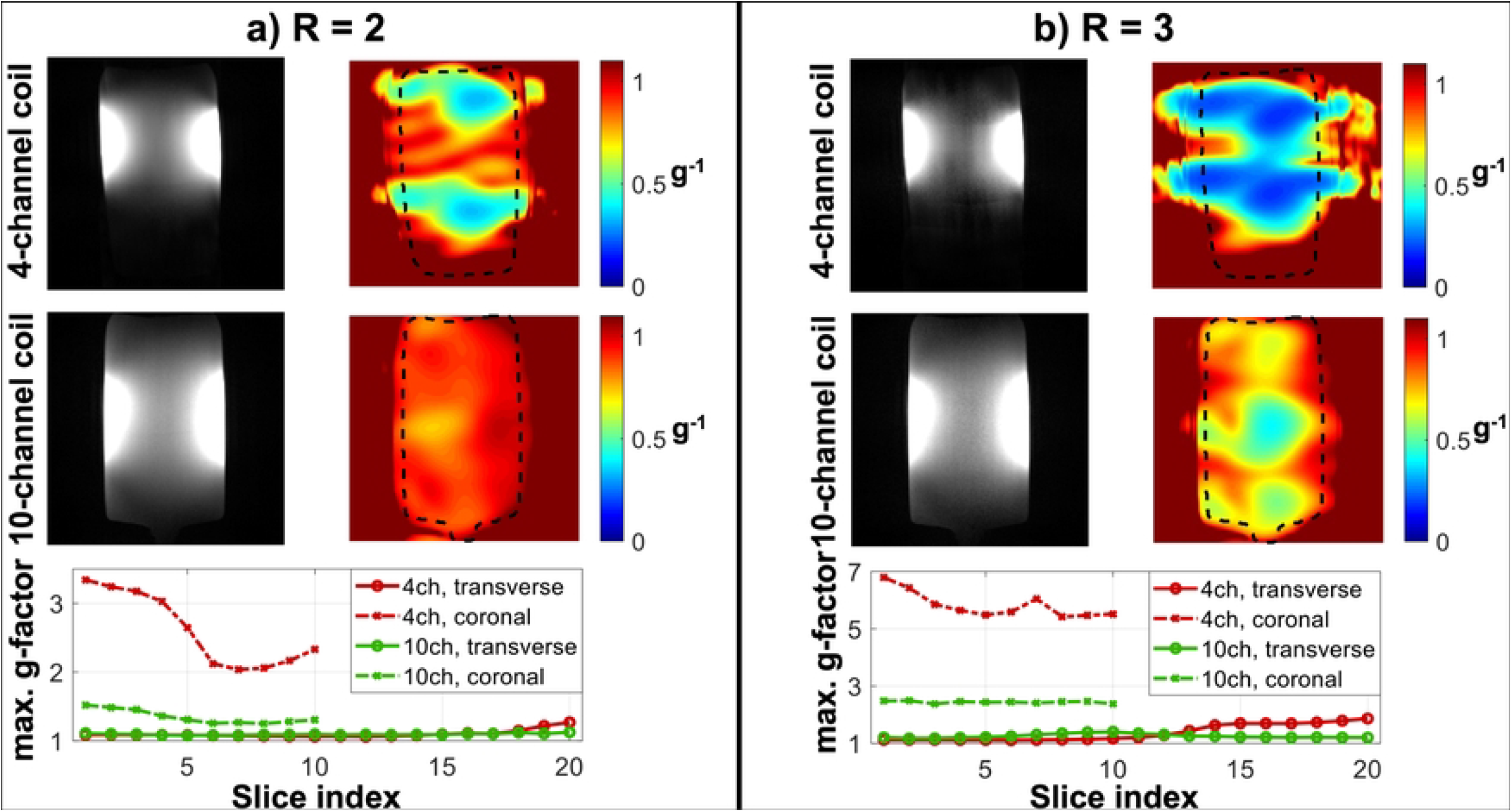
Estimated g-factor noise amplification in a phantom using the 4-channel and the 10-channel coils, at **(a)** R = 2 and **(b)** R = 3. Reconstructions as well as retained SNR (inverse g-factor) maps of a single coronal slice are shown for both coils and at both acceleration factors. The bottom row shows the maximum g-factor in each slice for both the transverse and coronal acquisitions. Note that the maximum g-factor values are shown using a different y-axis scaling in Figure (a) than in Figure (b).

### 3.2 *In vivo* comparisons

Figure 4 shows the estimated *in vivo* g-factor noise amplification using both coils at R = 2 (Figure 4a) and R = 3 (Figure 4b) for transverse acquisitions. The 10-channel coil achieves a small but significant (p = 0.003) g-factor reduction of 3 ± 3% at R = 2, and of 19 ± 9% at R = 3 (p < 0.001).

**Figure 4:**
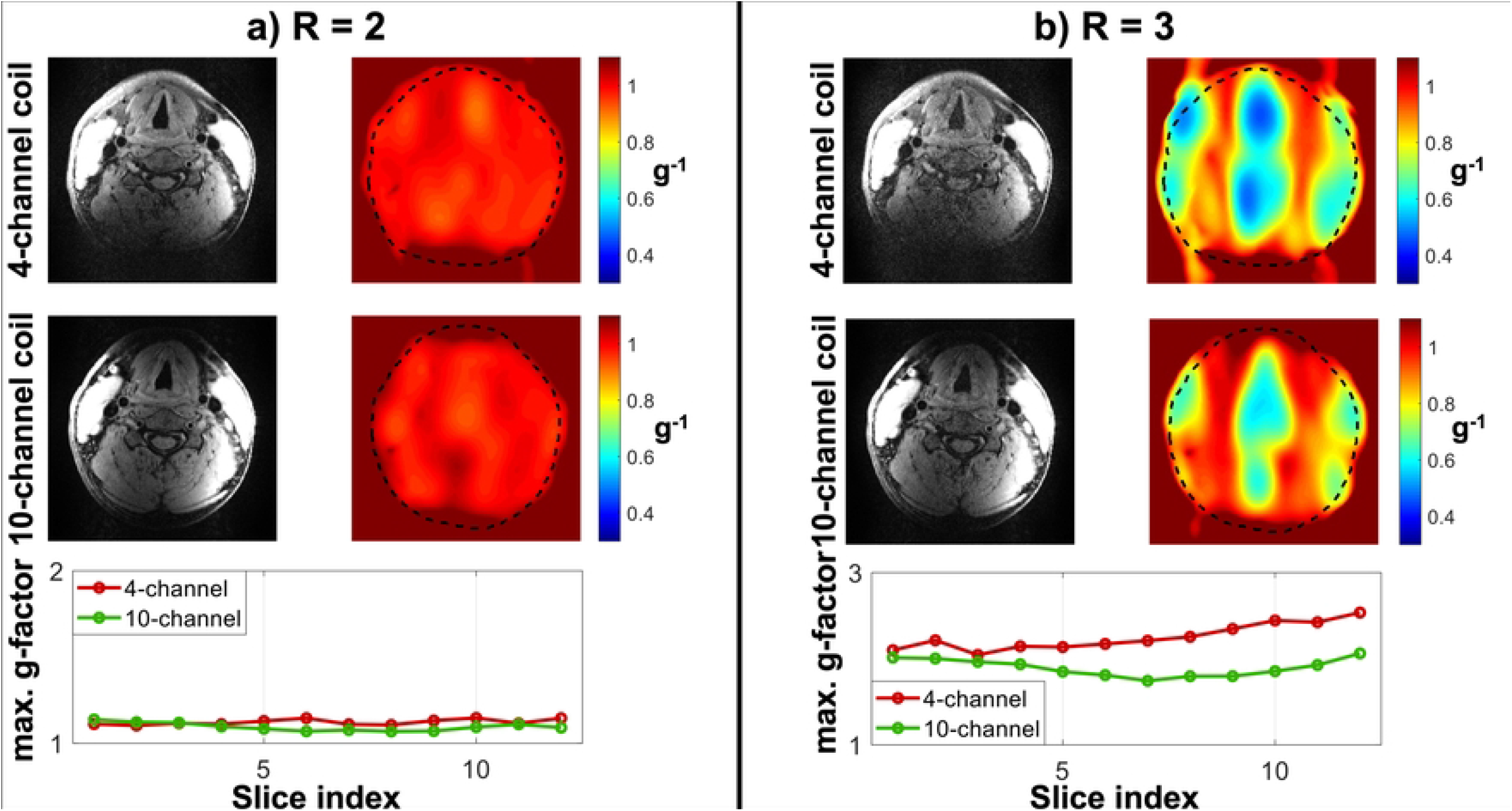
in vivo estimated g-factor noise amplification using the 4-channel and the 10-channel coils, at **(a)** R = 2 and **(b)** R = 3. All data is shown for transverse acquisitions. The top two rows show reconstructions as well as retained SNR (inverse g-factor) maps of a single transverse slice for both coils and at both acceleration factors. The bottom row shows the maximum transverse g-factor in each slice. Note that the maximum g-factor values are shown using a different y-axis scaling in Figure (a) than in Figure (b).

DANTE-MESE images at different echo times are shown in Figure 5 using both the 4-channel and the 10-channel carotid coil. The internal and external carotid arteries are clearly visible on the images from both coils at short echo times. Increased CNR using the 10-channel coil versus the 4-channel coil noticeably improves vessel visibility at longer echo times. The mean CNR between the vessel walls and the lumen are shown in Figure 5c for all subjects using both coils. The CNR is consistently significantly higher (+62 ± 11% for the 14 echo times; *p* p< 10^−5^at each individual echo time) when using the 10-channel coil, with the largest relative increases (up to +82%) at short echo times. In total, ∼14,000 vessel wall voxels were identified and compared across the 9 healthy volunteers.

**Figure 5:**
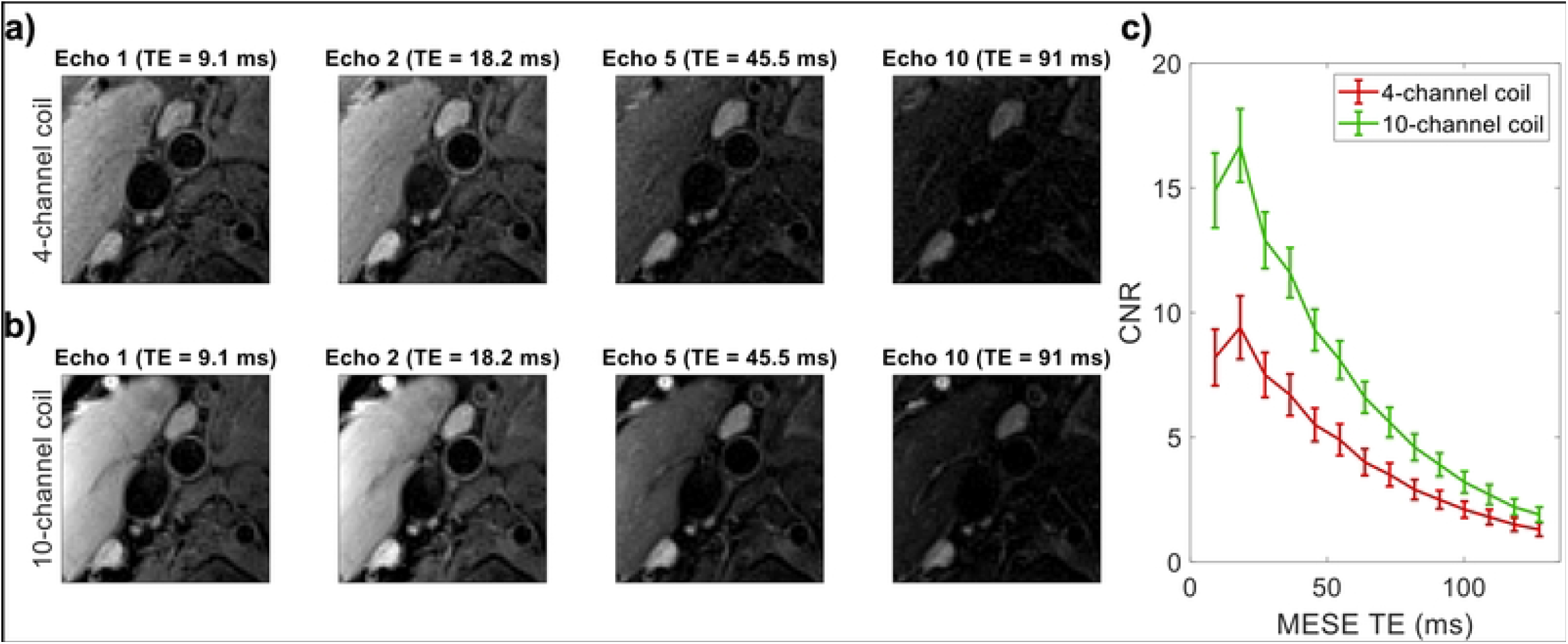
DANTE-MESE scans of nine healthy volunteers using both coils. All data is shown to the same greyscale. **(a-b)** Close-ups near the carotid bifurcation of a single volunteer at four different echo times using **(a)** the 4-channel coil and **(b)** the 10-channel coil. **(c)** Mean carotid wall/lumen CNR results of both coils across the nine volunteers.

The T2 value of healthy carotid wall tissue at 3T calculated for the 9 volunteers using the 14 echo times was 65.9 ± 14.1 *ms* (mean + SD) using the 4-channel coil, and 63.1 ± 13.6 *ms* using the 10-channel coil. The standard error of the T2 estimates was 13.6 ± 4.5 *ms* using the 4-channel coil, and 5.0 ± 3.3 *ms* using the 10-channel coil. The statistical distributions of data acquired with the 4-channel coil and 10-channel coil were significantly different for T2 values and standard errors (all tests rejected the null hypothesis with *p* p< 10^−5^). The improved SNR obtained with the 10-channel coil resulted in an average reduction of 24% on the standard errors of the estimated T2 values.

Figure 6 shows typical examples of DANTE-FSE T1-weighted images of the carotid arteries near the carotid bifurcation in two patients with atherosclerotic carotid artery disease, acquired using the 10-channel coil.

**Figure 6:**
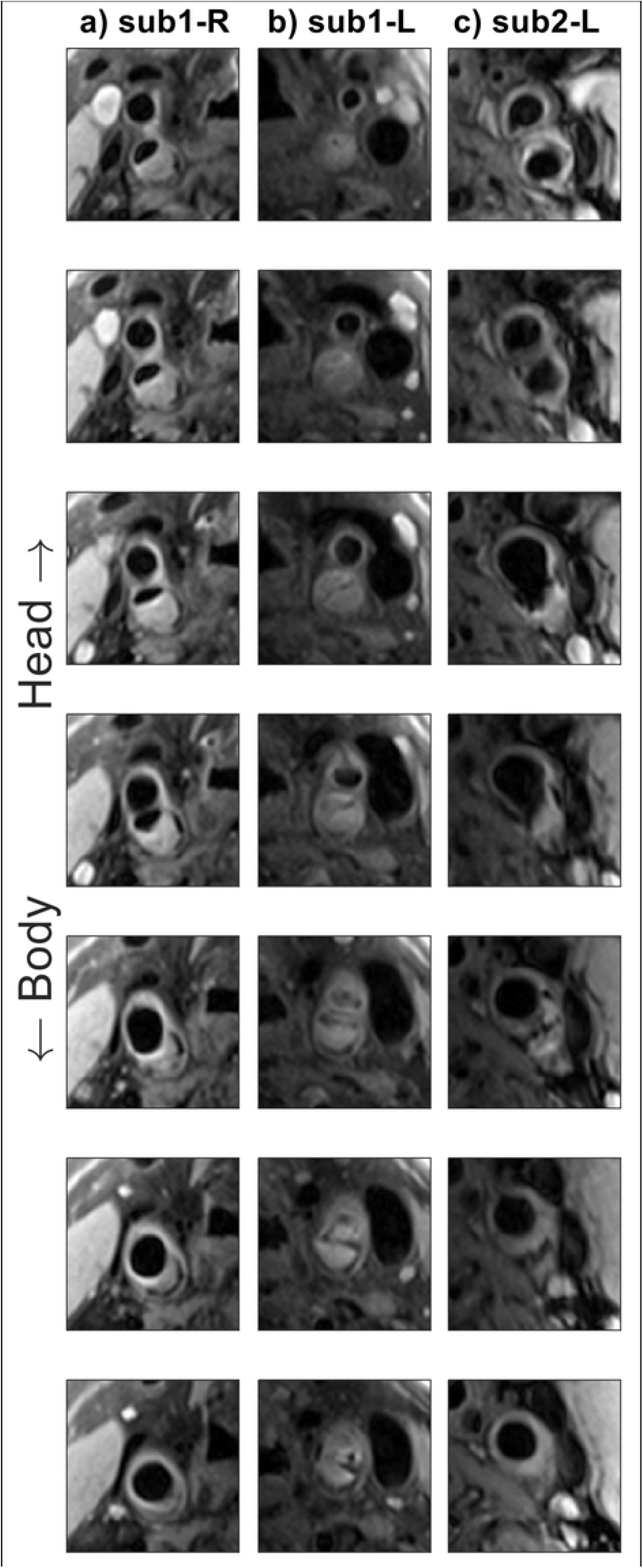
Consecutive DANTE-FSE T1-weighted slice segments showing the carotid bifurcations in two patients with atherosclerotic carotid artery disease, acquired using the new 10-channel coil. **(a-b)** Subject 1, left- and right-hand sides; **(c)** subject 2, right-hand side.

## 4. Discussion

In this study, a novel 10-channel phased-array coil design for carotid imaging at 3 Tesla was compared to a commercial 4-channel coil design. Data acquired in a phantom and in healthy volunteers were used to compare the SNR, vessel wall-lumen CNR, and parallel imaging noise amplification values of both coils. Additional patient data were included to show the typical image quality and carotid plaque details that can be achieved by the proposed coil.

The phantom results in Figure 2 Figure 3 show a significant increase in SNR (88 ± 2%) when using the proposed 10-channel coil compared to the commercial 4-channel coil, as well as a reduction in parallel imaging noise amplification for coronal acquisitions. Although the phantom data did not obtain a statistically significant reduction in noise amplification for transverse acquisitions, the *in vivo* results in Figure 4 did achieve significant reductions for transverse acquisitions.

The increase in SNR was largest close to the coils but consistently present throughout the phantom. In patients, the large increase in SNR near the edge of the neck is beneficial for imaging the relatively superficial carotid bifurcations. The consistent SNR improvement at greater depths indicates that this benefit can be maintained for patients with thicker necks or with atypical vasculature. Longitudinally, a mean increase in SNR of 88% was achieved over a distance of 10 cm in the phantom. Since the longitudinal position of the carotid bifurcation can vary by several centimetres between patients, this longitudinal consistency makes the SNR gain in the 10-channel coil beneficial to large groups of patients without requiring adjustments in coil positioning during a scan session. Away from the carotid bifurcation, the longitudinal SNR improvement, as demonstrated by the phantom data, provides improved coverage over a larger part of the carotid circulation.

In 2016, Hu et al.^23^ proposed an 8-channel carotid coil design, which they compared to the same commercial 4-channel coil design that is used for comparison in this paper. The proposed 10-channel coil obtained a 88% SNR increase in the phantom, consistently larger than the ∼40% increase achieved by Hu et al.^23^, slightly larger than the 70% SNR increase presented earlier for an 8-channel carotid coil relative to a custom-built 4-channel coil^15^, and similar to the SNR increase found when using the 16-channel coil proposed by Tate et al.^26^, which requires a larger number of receive channels and provides reduced positioning flexibility.

The lower g-factors when using the 10-channel coil (Figure 3) make it possible to visualize the carotid arteries at increased parallel imaging acceleration factors with limited noise amplification. This is especially important for cases when high-resolution data are acquired, such as for volumetric plaque quantification, or when multiple datasets with different contrasts need to be acquired for tissue characterisation, which would without additional acceleration require prohibitively long scan times. In practice, the minimal g-factor noise amplification using the proposed 10-channel design at R=2 means that data can be acquired with a substantial scan time reduction while maintaining clinical image quality. The large g-factor reduction in the longitudinal direction can be explained based on the difference in longitudinal position of some of the individual channels in the 10-channel coil, while all channels in the 4-channel coil are positioned in approximately the same longitudinal location.

The data acquired from healthy volunteers using the 10-channel coil and the 4-channel coil (Figure 5) show significant SNR and carotid wall-to-lumen CNR improvements which are consistent with the phantom studies. The mean CNR increase of +62% (up to +82% for the shortest TE) at the bifurcation is consistently larger than that achieved by the 6-, 8-, and 30-channel coil designs reported by Zhang et al.^20^, who measured their CNR increase relative to the same 4-channel coil used in this study, and with similar FOV and voxel size. ^26^Compared to the 4-channel coil, the increased CNR obtained using the 10-channel coil provided an improved vessel visibility around the carotid bifurcation at all echo times in the DANTE-MESE acquisitions, and a reduced error on the estimated T2 values of the healthy vessel wall across the 9 healthy volunteers.

In the patient data, as shown in Figure 6, the high CNR of the T1-weighted images provided by the 10-channel coil produced clearly visible and clinically useful vessel and plaque delineation over the length of the carotid bifurcation. The carotid bifurcation was clearly delineated for all 12 patients that were scanned using the 10-channel coil array, despite differences in longitudinal location of the bifurcations, benefiting from the increased longitudinal coverage of this proposed coil design (Figure 2b). In compliance with the approved ethics agreement, additional patient data using the 4-channel coil were not acquired and are therefore not available for comparison in this study.

Zhang et al. compared 4-, 6-, 8-, and 30-channel coils, and found that designs with fewer channels achieved higher SNR coverage than the 30-channel coil, while the 30-channel coil facilitated higher parallel imaging acceleration factors^20^. The 10-channel coil configuration proposed in this study offers reduced g-factor noise amplification in accelerated coronal acquisitions, while achieving increased SNR coverage compared to the commercial 4-channel coil and two previously presented 8-channel coils^15,23^ as well as improved CNR performance compared to the 6-, 8-, and 30-channel coil designs^20^. The proposed design can enable accurate imaging of the carotid bifurcation at high resolutions using multiple contrasts or quantitative mapping for plaque characterization within shorter scan times.

## 5. Conclusion

A novel 10-channel phased-array coil configuration achieved better visualization of the carotid bifurcation, with significantly increased SNR and CNR and decreased g-factor noise amplification. This design can facilitate improved characterization of atherosclerotic plaques in the carotid arteries within shorter scan times.

## Data Availability

All relevant data are within the manuscript and its Supporting Information files.

## Abbreviations

CNR: contrast-to-noise Ratio
DANTE: delay alternating with nutation for tailored Excitation
FSE: fast spin echo
g-factor: geometry factor
GRAPPA: generalized autocalibrating partially parallel acquisition
MESE: multi-echo spin echo
MRA: magnetic resonance angiography
MRI: magnetic resonance imaging
RF: radiofrequency
SD: standard deviation
SENSE: sensitivity encoding
SNR: signal-to-noise ratio
TOF: time-of-flight
TR/TE: repetition time/echo time

## Acknowledgements

We thank Peter Manley and Alison Fletcher for making it possible to acquire phantom data at the Acute Vascular Imaging Centre (AVIC) despite restrictions due to COVID-19, and Aaron T. Hess for valuable discussions during the preparation of this manuscript. The Wellcome Centre for Integrative Neuroimaging is supported by core funding from the Wellcome Trust (203139/Z/16/Z). MdB acknowledges studentship support from Siemens Healthineers and the Dunhill Medical Trust. PJ thanks the Dunhill Medical Trust and the NIHR Oxford Biomedical Research Centre for support. LB acknowledges support from the British Heart Foundation (PG/15/74/31747).

## Notes

### Competing Interest Statement

I have read the journal's policy and the authors of this manuscript have the following competing interests: MdB receives studentship support from Siemens Healthineers and CR is an employee of PulseTeq Limited.

### Author Declarations

Healthy volunteer data were acquired under an agreed technical development protocol approved by the Oxford University Clinical Trials and Research Governance office, in accordance with International Electrotechnical Commission and UK Health Protection Agency guidelines. For patient imaging, ethical approval was obtained from the UK National Research Ethics Services and patients provided written informed consent.

